# HLA-restricted Biology of Severe Cutaneous Adverse Reactions (SCARs): Dapsone-induced Hypersensitivity as an Example

**DOI:** 10.1101/2025.05.24.25326117

**Authors:** Divya RSJB Rana, Sarita Manandhar, Anjana Singh, Deanna Hagge, Jarina Joshi

**Author notes:** Corresponding author: Divya RSJB Rana, Deanna Hagge, Jarina Joshi.

## Abstract

HLA proteins are pivotal in antigen processing and adaptive immunity. This study investigates the differential expression of class I and II HLAs in human skin, focusing on HLA-B’s role in drug-induced hypersensitivity syndromes (SCARs). Using RNA expression data from the Human Protein Atlas, we identified HLA-B as the highest expressed class I protein in exposed and non-exposed skin, with significant gender– and age-related variations. Male individuals exhibited higher HLA expression in sun-exposed skin, particularly for HLA-B, which aligns with reported higher SCAR incidences in males. Expression levels increased with age, suggesting a potential link to the heightened SCAR prevalence in older populations. Additionally, molecular docking analyses were performed to investigate HLA alleles associated with dapsone hypersensitivity. Using SWISS DOCK and AutoDock Vina, we identified four non-HLA-B13:01 alleles—B13:13, B40:01, B40:10, and B15:25—with significant binding potential, potentially explaining hypersensitivity cases not attributed to HLA-B13:01. These findings offer insights into the molecular basis of hypersensitivity syndromes and the epidemiological variability across populations.

## Introduction

### Severe Cutaneous Adverse Reactions (SCARs)

Human SCARs which include drug-induced hypersensitivities or allergies, once regarded as idiosyncratic, have been repeatedly associated with specific types of Human Leukocyte Antigen (HLA) alleles as genetic risk factors (Chung 2007). The HLA molecules are highly polymorphic human cell-membrane proteins that help in adaptive immunity by processing endogenous or exogenous cytosolic proteins and presenting them to T cells for immune surveillance. High population variability of these proteins (collectively denoted as MHCs, major histocompatibility complexes in Gnathostomes) helps to ensure that at least a small proportion of individuals in the population are always able to present high avidity peptide epitopes of proteins of virulent organisms and stimulate the T cells (Fiorillo 2017). Over 20,000 class I and 10,000 class II HLA alleles have been reported (https://www.ebi.ac.uk/). These molecules have evolutionary relationships and have been grouped into bigger categories. “Supertypes” are the classification of individual HLA class I alleles into more broader categories based on how the B and F pockets of the peptide binding grooves (PBGs) in the HLAs interact with the peptides (Sette 1998, 1999). The class I HLAs are heterodimers of variable □ chain and conserved beta2-microglobulin proteins. The □1 and □2 domains, which are □ helices within the □ chain of class I HLAs, make up the PBG. Residues 7, 9, 24, 34, 45, 63, 66, 67, 70, and 99 make up the B, and residues 74, 77, 80, 81, 84, 95, 97, 114, 116, 123, 133, 143, 146, and 147 make up the F pockets (Nguyen 2022). Among them, 9, 45, 63, 66, 67, 70, and 99 are key residues for B, and 77, 80, 81, and 116 are key for F (Sidney 2008) pocket. The B and F pockets in combination with other pockets make an enclosed PBG on the exposed surface of class I MHC molecules. The supertypes are based on similarities of these residues playing roles in selecting peptides in their respective groves. There are predilections of HLAs for some specific peptides, specifically the second and last amino acids of the peptide fragments bound in the HLA class I grooves (Konstantinou 2017, Sidney 2008). These residues in the peptide fragments make stable bonds with residues surrounding the B and F pockets in the HLA class I PBGs. Stimulation of T cells depends on the binding of T cell receptors to the MHC complex which includes dimer of the MHC and antigenic peptide. This stimulation is mandatorily assisted by other co-stimulatory molecules for proper activation of T cells. Allergic stimulation of T cells in response to exogenous drugs makes use of this pathway and leads to surprisingly similar sets of symptoms (liver and skin are usually invariantly involved in all DRESS episodes) precipitated by these widely varied groups of drugs. Class I HLA-restricted SCARS have been explained based on three hypotheses: hapten/pre-hapten hypothesis, altered peptide repertoire hypothesis and pharmacological interaction hypothesis. More recently, two different approaches/hypotheses have been discussed to explain the “altered repertoire hypothesis” for the SCARs (Pichler 2013) (Figure 1). The first is that the drugs bind to the PBGs and change the internal structure of the groove leading to different kinds of peptides being bound to the HLAs. The other is changes in the orientations of the peptide ligands already bound to nascent HLA molecules. Both scenarios lead to stimulation of T cells. As peptide loading to MHCs is preceded by the selection of peptides by TAP proteins (Transporter of Antigen Processing) and there have been no reports of drugs binding toTAPs, there is a high chance that drug-PBG binding leads to a change in 3D orientation of peptides that are already bound to the PBGs (scenario two) and binding of entirely different peptide (scenario one) is less likely. This is also supported by experiments where glutaraldehyde fixation of proteasomal processing of peptides is unable to prevent drug stimulations (Ostrov 2012, Illing 2013, Zhao 2019). However, some in vitro studies do show the requirement of processing of the drugs in the proteasomal pathway for drug stimulations (Bell 2013, Zhao 2019), and thus the role of proteasomal processing can not be ruled out and further studies are required. The changes in the peptide presentation (altered peptide repertoire hypothesis) lead to “non-self” stimulation of T cells. A more recent hypothesis for drug hypersensitivity or SCARS is that the drug may bind to the HLA or T cell receptor directly and stimulate the T cell without any role of the peptide binding (p-i concept or pharmacological immune receptor interaction concept). Jiang et al 2022 showed HLA-B*13:01 with bound dapsone could stimulate T Cells in the absence of any peptides. Thus different mechanisms may play roles in the precipitation of the SCARs.

**Figure 1:**
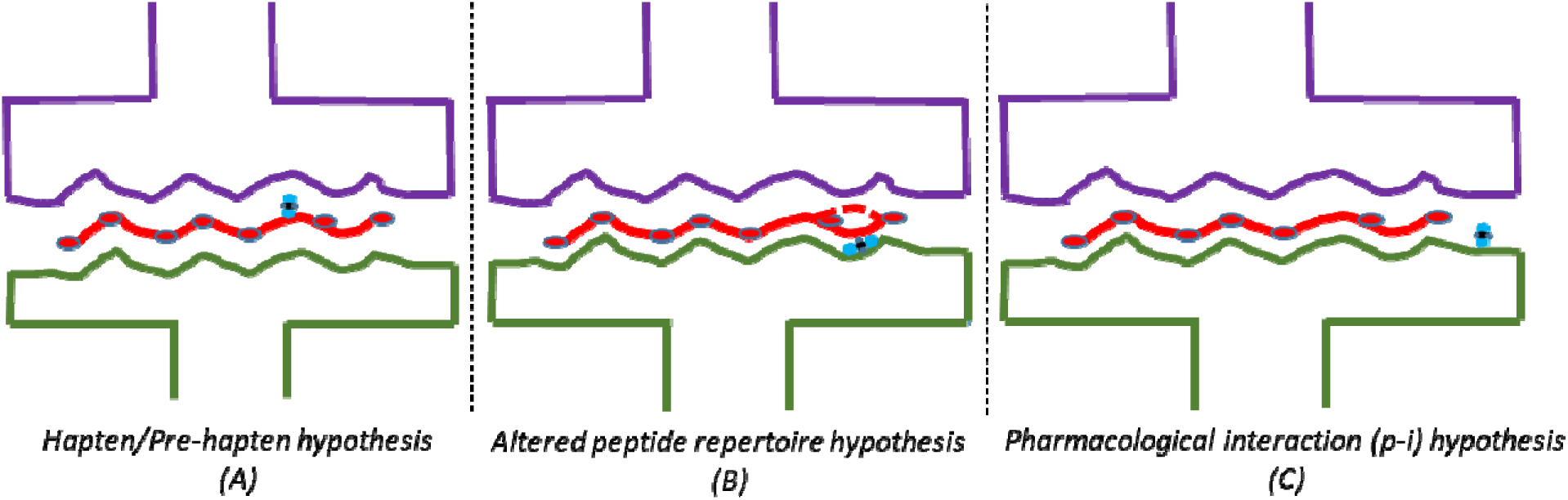
Hypothesized mechanisms of T cell stimulation by drugs in SCARs. A. changed/haptenated peptides which are antigenic/non-self; B. changes in usual peptide repertoire of specific HLAs by binding to peptide binding grooves; C. direct activation of T cells by HLA bearing APCs. Purple colored upper structure is the T cell receptor, the greenish structure below is HLA, the red beaded line is peptide with amino acid moieties, and the bluish structure is dapsone.

### Mechanism of SCAR/Drug Hypersensitivity for Dapsone

Dapsone is an antibiotic and anti-inflammatory drug that has been approved by the FDA for treatment of leprosy and dermatitis herpetiformis. Additionally, dapsone is also used for the treatment of *Pneumocystis jiroveci* pneumonia, Brown Recluse spider bite, malaria prophylaxis and different variants of autoinflammatory skin diseases like pyoderma gangrenosum, pemphigus vulgaris, IgA pemphigus, etc among others. Dapsone hypersensitivity (DHS) was first described in late 1940s (Lowe 1949) and more systematically studied by Richardus and Smith in 1989 resulting in a widely used DHS criteria called the “Richardus and Smith criteria”. In 2013, two independent groups in China discovered a human leukocyte antigen HLA-B*13:01 to be a significant risk factor for DHS. Since then many studies have validated this association. Watanabe 2017 did molecular docking studies on how dapsone may bind its known HLA risk allele HLA-B*13:01. They predicted the binding of dapsone to F pocket of HLA-B*13:01’s PBG. As close members of HLA-B*13:01 allele, such as HLA-B*13:02 and HLA-B*13:03, are not known to elicit any SCARs, and as these alleles only differ in 3 moieties (amino acids 118,119 and 121) in the F pocket of PBG, the predictions were plausible. Later molecular modeling by Jiang 2022 also showed that dapsone is bound into F pocket with hydrophobic interactions to 108Y, 109Y, 118I, 119I, 142Y, 147Y and 171W of HLA-B*13:01. They also carried in vitro studies to differentiate the roles of amino acid acids 118,119 and 121, thereby explaining why HLA-B*13:01 is a potent activator of T cells but HLA-B*13:02 is not.

In vitro studies by Jiang 2022 showed that two simultaneous mutations at 118, 119 or 121 could abrogate stimulation of T cells.

Many studies on dapsone allergy before the HLA association era were done by Craig K. Svensson’s group at Wayne State University. Then, N-hydroxylamine of dapsone was considered a major factor in drug allergy/hypersensitivity (Svensson 2003). Additionally, presence of N hydroxylation capacity in keratinocytes which are the primary target of hypersensitivity reaction, boosted the belief that N hydroxylamine had some role in causation of allergy (Khan 2007). Later Alzahrani (2018) and Zhao (2019) in the University of Liverpool, UK found the role of N-hydroxyl dapsone in initiation of allergic CD4 T cells. But the role of CD4 T cells is not clear when SCARs are known to be associated with class I HLAs which incite CD8 T cells. Most of the T cells activated after in vitro stimulation of PBMCs of DHS patients or in blister cells of DHS patients were CD8 T cells (Jiang et al 2022). The need for antigen processing was dispensable and direct “superficial” binding of dapsone to HLA could bring about DHS. The presence of CD4 T cells activation in PBMCs from patients with varied HLA backgrounds hint that only HLA-B*13:01 (or other similar HLA) bearing cells and CD8 T cells are major factors in activation of SCARs.

In spite of these, the association between HLA risk factor and SCARs do not seem to be perfect. A recent meta-analysis has found that ∼15% of DHS patients did not bear HLA-B*13:01 (Satapornpong 2021) and another ∼15% of non-bearers had DHS. DHS is considered a DRESS (Drug Reaction Eosinophilia and Systemic Syndrome) variant (Lorenz 2012) which is one of the many widely cutaneous drug adverse events that include MPE (maculopapular exanthema), FDE (fixed drug eruption), urticaria, SJS-TEN (Steven Johnson Syndrome and Toxic Epidermal Necrolysis), etc (Roujeau 2005). DRESS, as defined by the European Registry for Severe Cutaneous Adverse Reactions, is defined by a set of clinical and laboratory criteria (Kardaun et al 2013). The criteria match with those set by the Japanese consensus group (Shiohara 2012, 2019) who call DRESS as DiHS (Drug-induced Hypersensitivity Syndrome). DiHS only differs from DRESS by inclusion of a criteria for reactivation of herpesvirus during these hypersensitivity reactions. In spite of these, most of the clinical symptoms are common and these symptoms are present in DRESS/DiHS brought about by a variety of drugs. These hint that the pathology of DRESS/DiHS is similar in all drug reactions. How can we account for that? In this study, we try to perform molecular docking analyses for HLAs other than HLA-B*13:01 to effectively bind dapsone. Additionally we do in silico immunopeptidome analyses to see amino acid preferences in peptides that may bind to HLA-B*13:01 in keratinocytes and hepatocytes to bring about the HLA-T cell activation.

## Skin and liver symptoms

The central mechanism in drug-induced hypersensitivity or SCARs reaction is T cell activation. T cells are activated by their HLA-restricted biology. In the PBGs of HLAs, the subtle differences in peptides determine the fate of being self or non-self, and hence, the activation of T cells. One of the peculiar features of all drug-induced DRESS episodes is involvement of skin and liver. DRESS and/or SJS-TEN syndromes are defined by rashes or exfoliation of skin/muco-cutaneous layers and increased liver enzymes and bilirubins. In fact, mortality in drug-induced hypersensitivity or SCARs is predicted by Hy’s law which hypothesizes the higher odds of mortality based on increased values of transaminases and alkaline phosphatase (FDA 2009). This hints at some kind of common mechanism taking place in liver and epidermal cells. All nucleated cells bear the same class I HLAs in a given individual. If the proteomes of tissues are similar or overlapping, there is a chance of a similar or overlapping set of immune peptides (“immunopeptidome”) being loaded to the HLAs in the two tissues. In this study we used in silico method to test this hypothesis. Data from ProteinAtlas.org was used to study the impact on human peptidome.

## Methods

### Expressional Analyses using Protein Atlas Data

RNA expression data for human leukocyte antigens (class I and II) were transferred from Protein Atlas (proteinatlas.org) to excel and further analyses were done for expression based on age groups, sex, and skin (exposed skin vs unexposed skin (pubic)). Data were present in normalized transcripts per million transcripts (nTPM). Mann Whitney test was used to compare various groups as all the data were non-parametric.

#### Non-HLA-B*13:01 HLA alleles

Data were searched in the literature for HLA loci information at B locus in dapsone hypersensitivity (DHS) and control patients to extract any information about HLAs other than HLA-B*13:01 that could be responsible for DHS. Prevalence of such alleles in populations where association between HLA-B*13:01 and DHS were all retrieved from allelefrequencies.net (The Allele Frequency Net Database, AFND, Gonzalez-Galarza 2018). Simple/rough Odds Ratio (OR) were calculated based on all available data.

#### Molecular Docking

Molecular Docking were performed by SWISS Dock and AutoDock Vina for non-HLA-B*13:01 HLA alleles found to be reported in DHS cases. DNA and protein sequences for required HLA-B alleles were obtained from IPD-IMGT/HLA database at www.ebi.ac.uk (Barker 2023). To build the homology models of the non-HLA-B*13:01 alleles, templates for models were searched both in NCBI-BLAST and online model builder SWISS MODEL (Waterhouse 2018, swissmodel.expasy.org). Homology modeling by SWISS MODEL provided ratings of different targets based on GMQE and sequence identity. These criteria along with others, including sequence coverage and resolution, were considered for our decisions to finally select for model building. Models with better resolutions were considered better. Even if the coverage was low, if the homology/identity was high, those were chosen for modeling. Few of the selected models had peptides. They were removed by AutoDock before uploading to SWISSDock. Necessary preparations for HLA models (removing ions or other elements, adding polar hydrogen, removing water, adding Kollman charges, etc) were done in AutoDock before using the pdbqt file for molecular docking.

For AutoDock Vina, the grid was selected in a small space of 27000 cubic Angstroms with exhaustiveness of 32 (Trott 2010).

### Immunopeptidome analyses

Immunopeptidomic analyses were carried to find major amino acids in 9-amino acid peptides that bind to F pocket of peptide binding groove of HLA-B*13:01. As the dapsone is thought to bind the F pocket (Watanabe 2017, Satapornpong 2020), it is expected that the amino acid at F pocket should be less bulky when dapsone is already bound. Using available free resources like NetMHCPan4.1 (Reynisson 2020), it is difficult to predict peptide sequences that bind to HLAs in a state when a drug is bound. But we tried to find if such less bulky amino acids are present in “strong binding” peptides as these could act as “back up” peptides whenever there dapsone bound to F pockets. To search for binding peptides, we needed proteins. First of all, IHC (immunohistochemistry) protein expression data for humans (HPA IHC data) were downloaded from Protein Atlas database (Sjöstedt 2020, The Protein Atlas Database, 2024: proteinatlas.org). Based on “Level” of expression and “Reliability” of expressional values provided, proteins expressed at “high” levels and tagged as of “approved” or “enhanced” reliability were selected. Further only those proteins in individual tissues which were common to proteins in Keratinocytes of skin 1 (sun-exposed) (total 458 proteins in Keratinocytes) were selected for immunopeptidome analysis. The protein symbols of the common proteins of each tissues/cell types were used to find protein sequences from Uniprot (The Uniprot Consortium, 2023) using the “Batch Retrieval and ID Mapping function”. Specifically, “ID Mapping” Tool was selected and the uploaded protein symbol data (.txt file) was entered as Gene Name of UniProt database to UniProtKB database. Our hypothesis for the occurrence of liver and skin symptoms during SCARs was the possibility of similarity in proteins expressed in these two tissues, hence similar processed HLA peptides, present in these two kinds of tissues, coupled with high HLA-B expression. The “reviewed” protein sequences from human (Homo sapiens) source in FASTA format obtained from UniProt were uploaded to NetMHCpan4.1 database (<u>NetMHCpan 4.1 –</u> <u>DTU Health Tech – Bioinformatic Services</u>) to retrieve 9 amino acid long peptides that would bind (“strong” and “weak” binders with EL ranks <0.5 and 0.5 to 2% respectively) HLA-B*13:01. EL rank meant Eluted Ligand ranks which not only represented the binding affinity of the peptide but also processivity through the proteasomal pathway. Lower EL score represented higher binding and processive probability for presentation by the HLAs. Due to high number of binders, only those with EL ranks of 0.1 or less were used for generating sequence logo by Seq2logo (https://services.healthtech.dtu.dk/services/Seq2Logo-2.0/) using Shanon method, Hobohm1 clustering (threshold 0.63), weight on pseudocount 200.

Gene Ontology (GO), KEGG and Reactome analyses: The general motive for these analyses were to identify reasons behind why liver and skin are generally involved in all kinds of drug allergies. Special bioinformatics tools were used to identify the similarity between these tissues (hepatocytes and keratinocytes). All proteins from around 75 tissues retrieved from Protein Atlas with “high” expression and tagged reliability as “enhanced” or “approved” were used for analyses. GO (Gene Ontology) helps to identify functional roles of gene sets. Kyoto Encyclopedia of Genes and Genomes (KEGG) analysis links genes to higher level cellular and systemic processes focussing on metabolic pathways, signalling pathways and human diseases. Reactome pathway analysis focuses on molecular events and is more detailed for signalling and molecular pathways compared to KEGG. All three analyses were done using R package “clusterProfiler” and databases “org.Hs.eg.db” and “GO.db” using package manager “BiocManager” (Wu 2021). R codes were generated using conversation with ChatGPT (chatgpt.com).

## Results and Discussions

### HLA Protein Expressions

HLAs are the most polymorphic protein in humans and are directly responsible for antigen processing and thus, generation of adaptive immunity. These genes are responsible for the highest number of disease associations by genetic analyses (Fiorillo 2017). Unlike class II HLAs which process exogenous proteins for peptide presentation, class I HLAs process endogenous peptides. There are 3 subtypes for class I HLAs: HLA-A, HLA-B and HLA-C. HLA-B is the most frequently reported class I antigen with respect to SCARs association (Pavlos 2012, Negrin 2017). Thus a fundamental question remains on why HLA-B is preferred compared to other class I loci. We downloaded RNA expression data for HLAs from normal skin from Human Protein Atlas Database (proteinatlas.org) and compared the expression of different class I and II proteins. While analyzing the expressional data for each of the HLAs (class I and II), it was found that class I HLA had the highest expressions in both exposed and non-exposed human skin. Among class I HLAs, HLA-B showed the highest expression. Median expression for HLA-B, A and C in exposed skin were: 755.9, 549.3 (p<0.0001) and 407.7 (p<0.0001) in nTPM units (Figure 2 below). The highest expressional level for HLA-B could be the region why HLA-B is usually implicated in the majority of HLA-SCAR associations. When gender was used as a variable for differential expression, there was a trend of higher HLA expression in males compared to females. While there were no expressional differences in males and females for any of the class I HLAs in unexposed skin, males expressed significantly higher levels of HLAs compared to females in sun-exposed skin for HLA-A (534.2 vs 505.1, p<0.05) and HLA-B (732.6 vs 670.6, p<0.01). There were no differences for HLA-C in exposed skin (382 vs 377.3, p=ns). Males have been reported to have significantly higher drug-induced hypersensitivity/SCARs incidences (Kvedariene 2019, Lorenz 2012, Pagani et al 2022). When data for HLA-B expression in exposed skin were analyzed by age groups, it was found that the expression of HLA was in increasing trend with age. Compared to expression in 20-29 years age group (median expression 684.7 nTPM), the expression was higher but statistically insignificant in age group 30-39 years (median 698.3 nTMP, p=ns) and 40-49 years (median 736.5 nTPM, p=ns) and 70-79 years (median 764 nTPM, p=ns) age groups. But expression was significantly higher in 50-59 years (median 783.5 nTPM, p<0.05) and 60-69 years (median 799, p<0.01) age groups. Kvedariene 2019 reported higher prevalence of SCARs in elderly people. The proportion of people with dapsone hypersensitivity was higher for >=27 years than for <27 years age group (Lorenz et al 2012). Also Pagani et al 2022, reported higher prevalence of SCARs in people aged 18-65 years compared to those aged 12-17 years. Thus the higher expression of HLA-B in higher age groups could be one, if not sole, factor for increased incidence of SCARs in adult people. These data hint that the expressional level of class I loci in skin could be the reason for epidemiological variability of hypersensitivity prevalence in the population with respect to gender and age.

**Figure 2:**
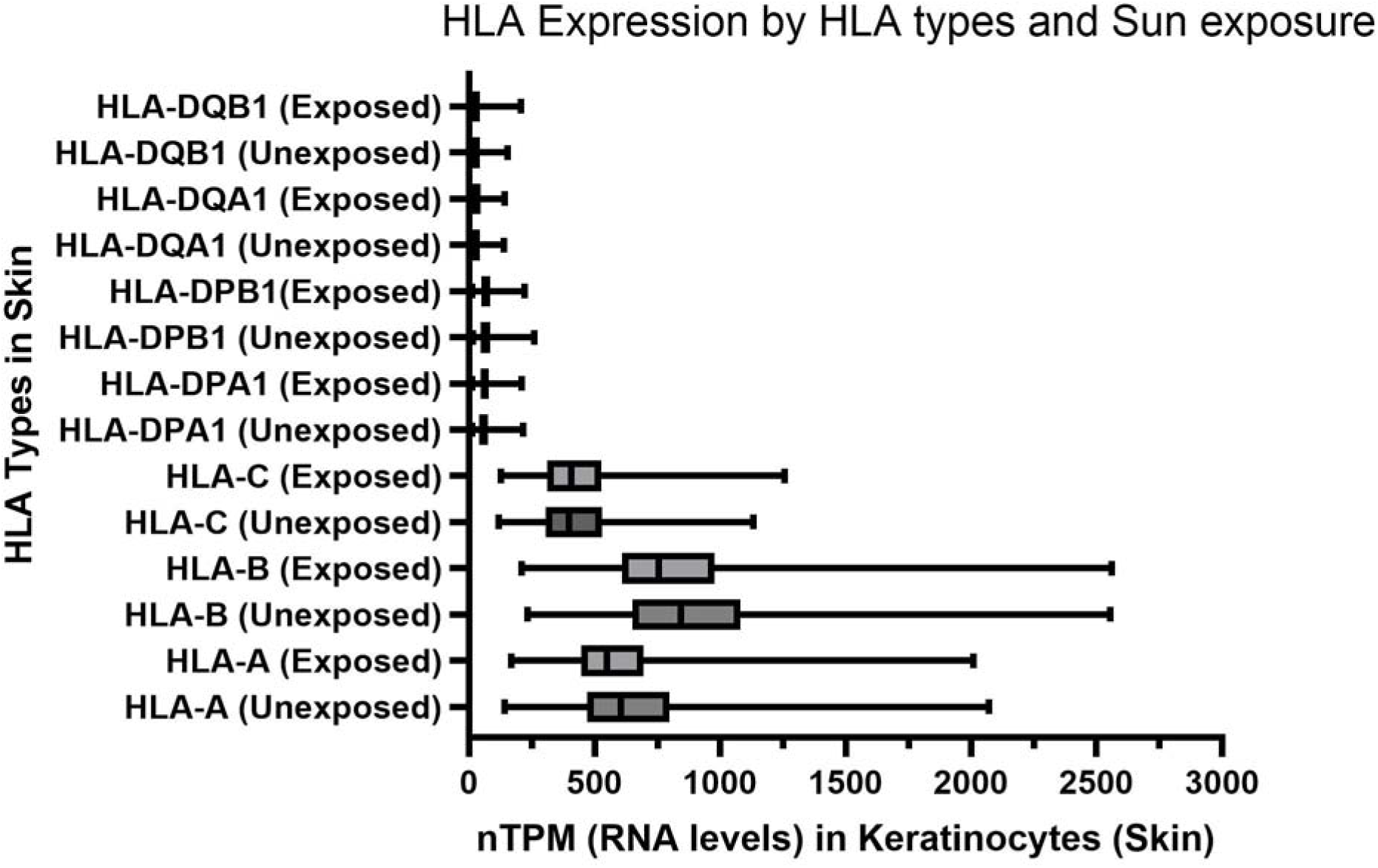
Normalized Transcripts Per Million reads data for HLA expression in exposed and unexposed human skin.

### Non-HLA-B*1301 in causation of DHS

The association between HLA-B*13:01 and DHS (dapsone-induced hypersensitivity) is well-known and validated in multiple countries (Wang 2013, Zhang 2013, Mustamu 2020, Chiramel 2019) but the presence of DHS in a small number of non-HLA-B*13:01 individuals are unexplained (allergic non-carriers). Data from literature were used to search for HLAs other than HLA-B*13:01 that caused DHS. Three articles reported individual HLA profiles in DHS cases (Wang 2013, Mustamu 2020 and Park 2020) and thus HLA-B alleles, other than HLA-B*13:01, implicated in DHS cases were short-listed for further analyses (Table 1). As shown in the table below, 4 alleles had significant odds ratio (OR): HLA-B*13:13 (OR 66.6 p=0.003), B*40:48 (OR 66.6, p=0.003), B*67:01 (OR Infinity, p=0.038) and B*40:10 (OR 28, p=0.074). The frequencies of the respective alleles in countries where HLA-B*13:01 and DHS associations have been established are also mentioned in the table (frequency data from allelefrequencies.net database, AFND, Gonzalez-Galarza 2018). Without laboratory evidence, it is hard to implicate the association conclusively. Further, statitical analyses could be affected by the presence of two HLA alleles (one significant and other insignificant) in the DHS cases and separate biological analyses for drug affinity for each of the allele need to be established before coming to any conclusion. For this, we carried in silico analyses by performing blind (SWISSDOCK) or targetted (AutoDock Vina) docking of dapsone into all HLAs (listed in table below, with significant or insignificant ORs) reported to be present in DHS cases.

**Table 1:** Alleles, other than HLA-B*13:01, that have been reported in known DHS cases with odds ratio (OR) for the association with DHS and frequencies in countries where HLA-B*13:01 and DHS associations have been established.

| Prevalence by allele frequencies.net |  |  |  |  |  | non-B*13:01 DHS alleles |  | DHS Cases |  | DHS controls |  | Statistical Parameters |  |  |  |
| --- | --- | --- | --- | --- | --- | --- | --- | --- | --- | --- | --- | --- | --- | --- | --- |
| India | China | Indonesia | Thailand | Taiwan | S Korea |  |  | Positive | Total | Positive | Total | OR | Sensitivity | Specificity | p-value |
| 2.2-24% | 1.8-42.2% | 2.6-3% | 4.2-8.2% | 0-45.1% | 0.042 | B*13:13 |  | 2 | 6 | 1 | 154 | 66.8 | 33 | 99 | 0.003 |
| 0-8% | 0-16.6% | 0.106 | NA | 2-22% | 1.6-2.2% | B*38:02 |  | 1 | 6 | 16 | 154 | 1.7 | 17 | 90 | 0.496 |
| NA | 0 | NA | NA | NA | NA | B*40:48 |  | 2 | 6 | 1 | 154 | 66.8 | 33 | 99 | 0.003 |
| 0-13.2% | 0-29.8% | 0.072 | 11-17% | 6-55.1% | 7.6-8.9% | B*40:01 |  | 2 | 6 | 35 | 154 | 1.7 | 33 | 77 | 0.623 |
| NA | 0-3% | NA | NA | 0.02-3.6% | 1.8-2.1% | B*67:01 |  | 1 | 6 | 0 | 154 | Infinity | 17 | 100 | 0.038 |
| 0.08-1.1 | 0-25% | 11.2-18% | 0.6-2% | 0-1.8% | 0-0.02% | B*35:05 |  | 1 | 6 | 4 | 154 | 7.3 | 17 | 97 | 0.176 |
| 0.06 | 0-3.2% | 12.4-22.2% | 1.4-2% | 0-1.8% | 0 | B*15:21 |  | 1 | 6 | 16 | 154 | 1.7 | 17 | 90 | 0.496 |
| 1.52-2.8% | 0-17.4% | 0-3.4% | 4-5% | 0-80% | 0 | B*15:25 |  | 1 | 6 | 2 | 154 | 14.4 | 17 | 99 | 0.109 |
| NA | 0-0.4% | NA | 0.308 | 0 | 0 | B*40:10 |  | 1 | 6 | 1 | 154 | 28 | 17 | 99 | 0.074 |

As mentioned in the methods section, appropriate homologs of the non-HLA-B*13:01 alleles were searched and built using NCBI BLAST and swissmodel.expasy.org. Out of the first 10 models, the best two were used for further analyses (better similarity in sequences, better Ramachandran plots, better resolutions). Models were also built for HLA-B*13:01 (known binder) and HLA-B*13:02 (known non-binder) to validate the binding by molecular docking. One of the models during the homology prediction was 7YG3.1.A which was recently constructed as a HLA-B*13:01 3D structure itself by Jiang 2022. 7YG3.1.A was one of our selected models for HLA-B*13:01, B*13:02 and B*13:13. After building the models (2 per allele), docking was performed in both SWISS DOCK (2 models per allele) and AutoDock Vina (one model per allele). Only docking with effective binding in the last pocket (in or near F pocket) was considered significant.

Both SWISSDOCK and AutoDock VINA could differentiate binding of dapsone to F pocket of HLA-B*13:01 (a known binder) and HLA-B*13:02 (known non-binder). Thus, we assumed that our method of docking is reliable. In this study, we only examined (visually) if the binding of dapsone ever took in the F pocket (farthest in the PBG) and if the binding affinity for the F pocket was sufficiently stable. Stability was decided based on better binding affinity compared to other binding poses in a given docking experiment. For SWISSDOCK results, if at least one homology model showed stable binding, then the binding was considered relevant.

SWISSDOCK found better binding stability at F pocket for B*13:01, B13:13, B*15:21, B*15:25, B*40:01, B*40:10 and B*67:01; intermediate for B*35:05 and B*40:48; and none for B*13:02 and B*38:02. AutoDock VINA showed (Figure 5) dapsone binding in F pocket stable for B*13:01, B*13:13, B*15:25, B*40:01 and B*40:10, intermediate stability for B*15:21 and B*40:48, and no binding for B*13:02, B*35:05, B*38:02 and B*67:01. With respect to epidemiological statistics (except for HLA-B*13:01 and B*13:02), significant association was for B*13:13, B*40:48 and B*67:01, better OR but insignificant or near significance for B*15:25 (p=0.07) and B*40:10 (p=0.1); and insignificant and lower OR for B*38:02, B*40:01, B*35:05 and B*15:25. As mentioned above, if we look at individual HLA-B* profile (Individual Case Table 3), B*13:13 and B*40:48 are together in case 1 and 2. As B*40:48 binding by SWISSDOCK and AutoDock VINA are of intermediate significance, B*40:48 may only be confounding in the epidemiological analyses. B*40:01 and B*40:10 binding by AutoDock and SWISSDOCK are among highly significant, though epidemiology shows elsewise. We considered these two alleles, B*40:01 and B*40:10, as significant binders as other factors (like plasma drug concentrations, etc.) may have confounded our epidemiological analyses. B*38:02, B*35:05 and B*67:01 (patient SN 3, 5 and 6 in Table 3) come along with B*40:01 and B40:10 and have low binding (B*67:01 have significant binding by SWISSDOCK but no F pocket binding by AutoDock Vina) and epidemiological association, thus we consider them as non-binders. Finally, one DHS case (without HLA-B*13:01) had B*15:21 and B*15:25 at B locus. As compared to B*15:21, B*15:25 has significant binding by SWISSDOCK and Autodock VINA aided by mediate statistical significance by epidemiology. Based on binding affinity, we consider B*15:21 as possible binder. Together, B*13:13, B*40:01, B*40:10 and B*15:25 may explain the causation of DHS in all six non-HLA-B*13:01 cases though we have not analyzed other immunological or biochemical factors that may concomitantly come into play. The cross-section of F pockets of selected alleles are shown below.

**Figure 3:**
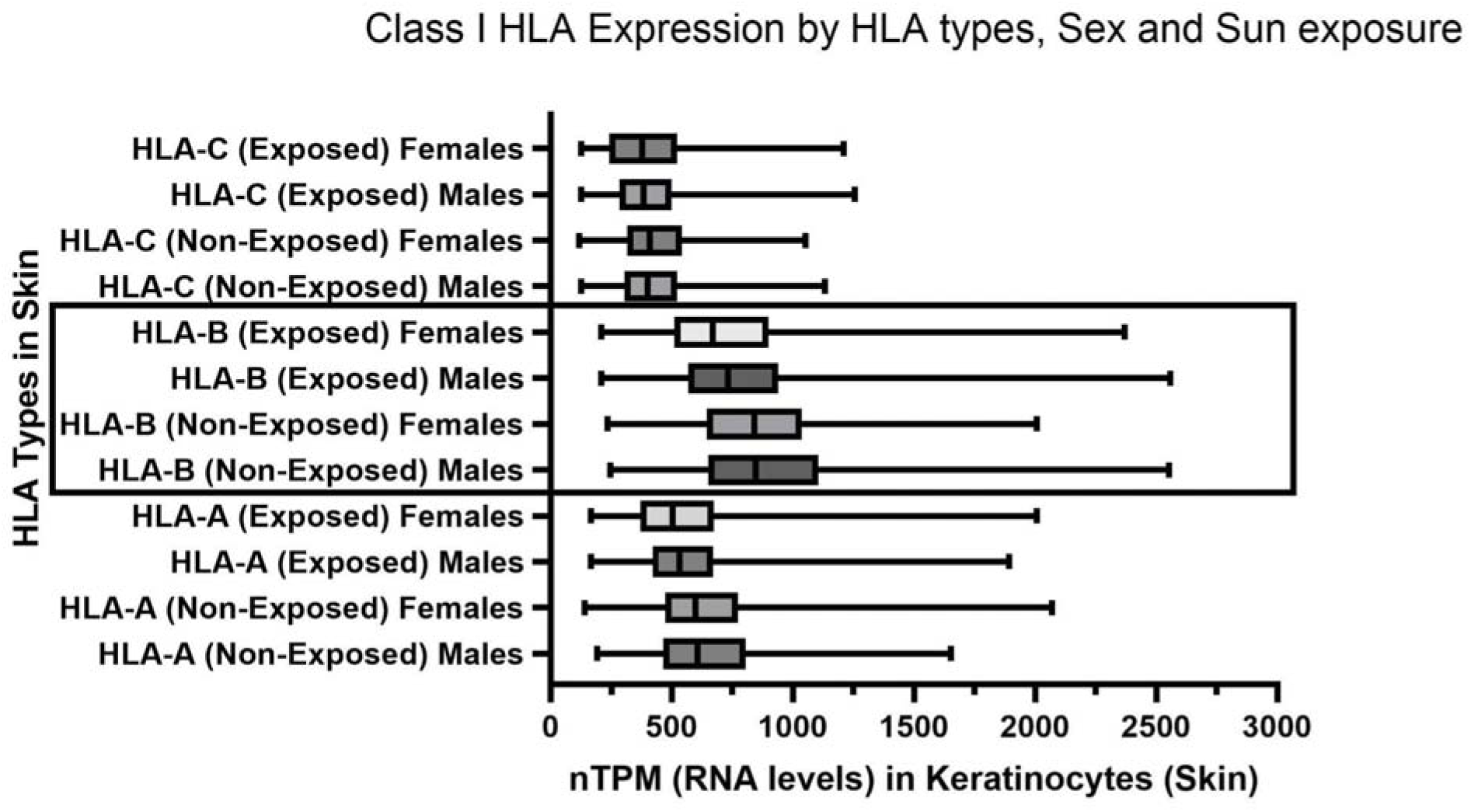
Normalized Transcripts Per Million reads data for class I HLAs expression in exposed and unexposed human skin with respect to males and females.

**Figure 4:**
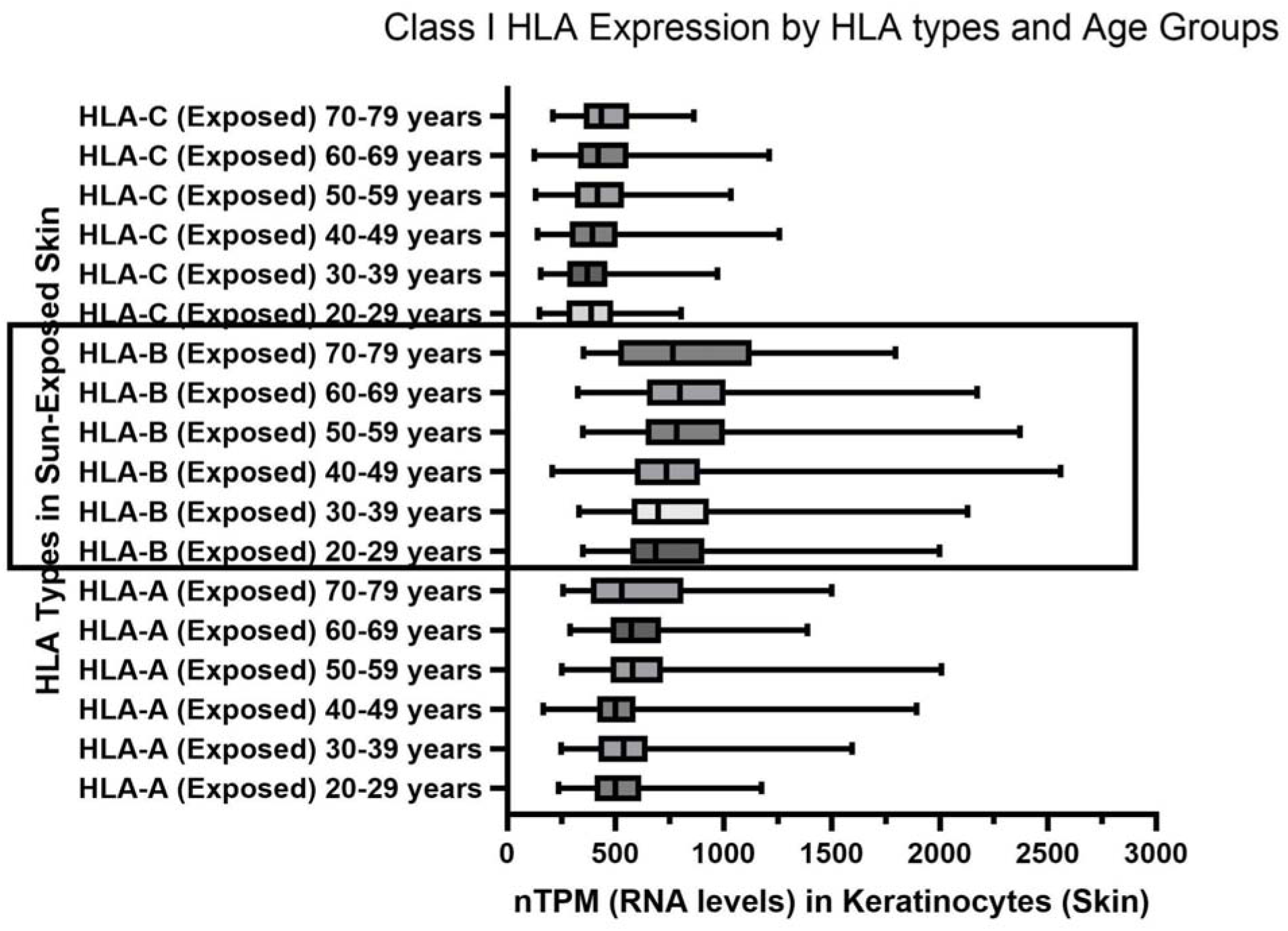
Normalized Transcripts Per Million reads data for HLA-B expression in exposed human skin for HLA-B in different age groups.

**Figure 5:**
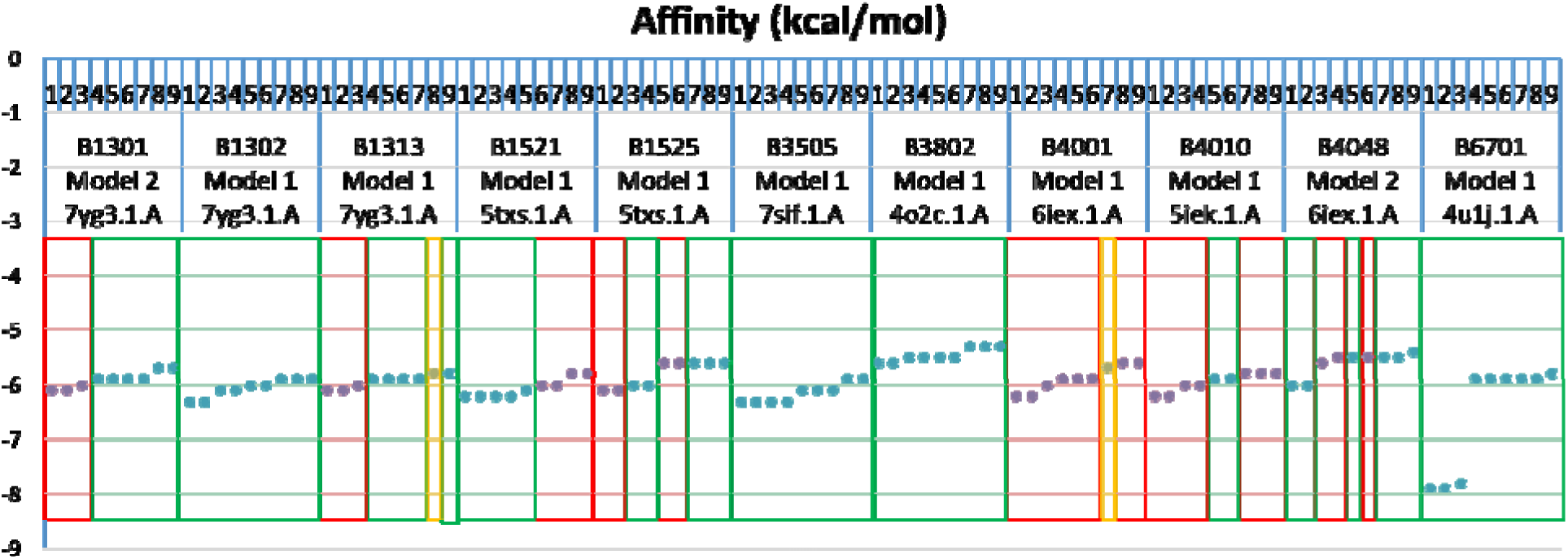
Results of AutoDock VINA for affinity of various poses of dapsone to different regions in the HLAs (see text). Red shade denotes binding to the F pocket. Blue shade denotes non-F peptide binding groove. Yellow shade denotes non-peptide binding groove. Number in the top represents 9 poses each for each docking experiment done in Autodock VINA.

**Figure 6:**
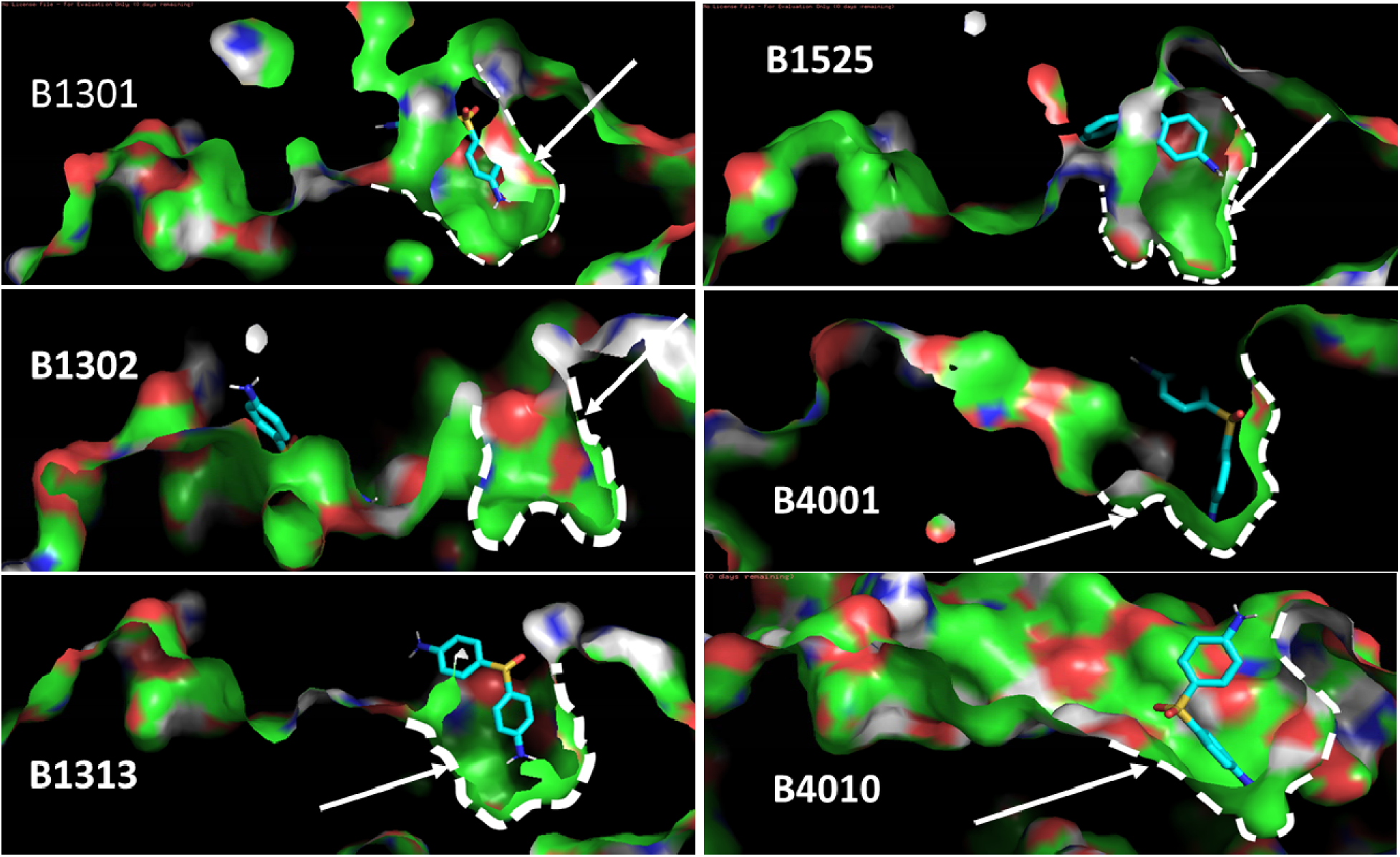
Binding of dapsone into F pockets of suggested non-HLA-B*13:01 HLA alleles. Pymol was used to identify the cross-section of the F pocket with files obtained from AutoDock Vina experiments.

**Table 2:** Summary of results from SWISSDOCK and AutoDock (AD) Vina and epidemiological statistics (see text).

| Method | Better stability at F or better Odds Ratio by epidemiology* | Intermediate stability at F or intermediate Odds Ratio by epidemiology | No binding at F or insignificant Odds Ratio by epidemiology |
| --- | --- | --- | --- |
| SWISSDOCK | B*13:01, B13:13, B*15:21, B*15:25, B*40:01, B*40:10, B*67:01 | B*35:05, B*40:48 | B*38:02, B*13:02 |
| AD VINA | B*13:01, B*13:13, B*15:21, B*40:01, B*40:10 | B*15:21, B*40:48 | B*13:02, B*38:02, B*35:05, B*67:01 |
| Epidemiology | B*13:13, B*40:48, B*67:01 | B*15:25, B*40:10 | B*38:02, B*40:01, B*35:05, B*15:21 |
| SUMMARY | **Binders: B\*13:01, B\*13:13, B\*40:01, B\*40:10, B\*15:25**<br>Non-binders: B*13:02, B*40:48, B*38:02, B*15:21, B*35:05, B*67:01 |  |  |

\*Epidemiology here refers to odds ratio calculated retrospectively, as in Table 1 above.

**Table 3:** All DHS cases without HLA-B*13:01 with reported non-HLA-B*13:01 alleles.

| SN | STUDY | HLA- A1 | HLA- A2 | HLA- B1 | HLA- B2 | HLA- C1 | HLA- C2 | Country |
| --- | --- | --- | --- | --- | --- | --- | --- | --- |
| 1 | Wang 2013 | A*24:02:01 | - | B*13:13 | B*40:48 | C*03:04:01 | - | China |
| 2 | Wang 2013 | - | - | B*13:13 | B*40:48 | C*03:04 | C*07:02 | China |
| 3 | Mustami: 2020 | NA | NA | B*38:02 | B*40:01 | NA | NA | Papua, Indonesia |
| 4 | Mustami: 2020 | NA | NA | B*15:21 | B*15:25 | NA | NA | Papua, Indonesia |
| 5 | Mustami: 2020 | NA | NA | B*35:05 | B*40:10 | NA | NA | Papua, Indonesia |
| 6 | Park 2020 | NA | NA | B*40:01 | B*67:01 | NA | NA | South Korea |

### Immunopeptidome analysis

#### Expressional analyses

Skin and liver injuries are very common features of drug hypersensitivity reactions. We hypothesized that similarity in the protein expressed in liver and keratinocytes could give rise to similar peptides in the HLAs and thus concomitant killing of skin and liver cells by activated T cells during the hypersensitivity.This could be the reason behind the concomitant injury of skin and liver during various kinds of drug allergies. We compared highly expressed proteins in different tissue with those proteins in keratinocytes. The total number of highly expressed proteins in keratinocytes was 458. Percent and numbers were calculated. Data were analyzed for top tissues/cell types with at least 60 proteins (arbitrarily chosen) in common with keratinocytes (25 tissues selected). In general, cell types with highest numbers of proteins expressed had higher expressional commonality with keratinocytes. This was depicted by a logarithmic linear equation: y = 108.93ln(x) – 541.96 (where x = number of highly expressed proteins for specific tissue and y = number of common proteins) with R² value of 0.6952. Contrary to our hypothesis, the commonality of expressed proteins between liver/hepatocytes and keratinocytes deviated (decreased) highest (–109.4%) with respect to what was estimated by the equation. Other deviations (increased) were seen for vaginal squamous epithelial cells (+35.7%) and cervix squamous epithelial cells (+35.9%). The higher commonality than expected could be due to the “epithelial” nature of the cell types. This hints at the highly non-epithelial nature of the hepatocytes.

**Figure 7:**
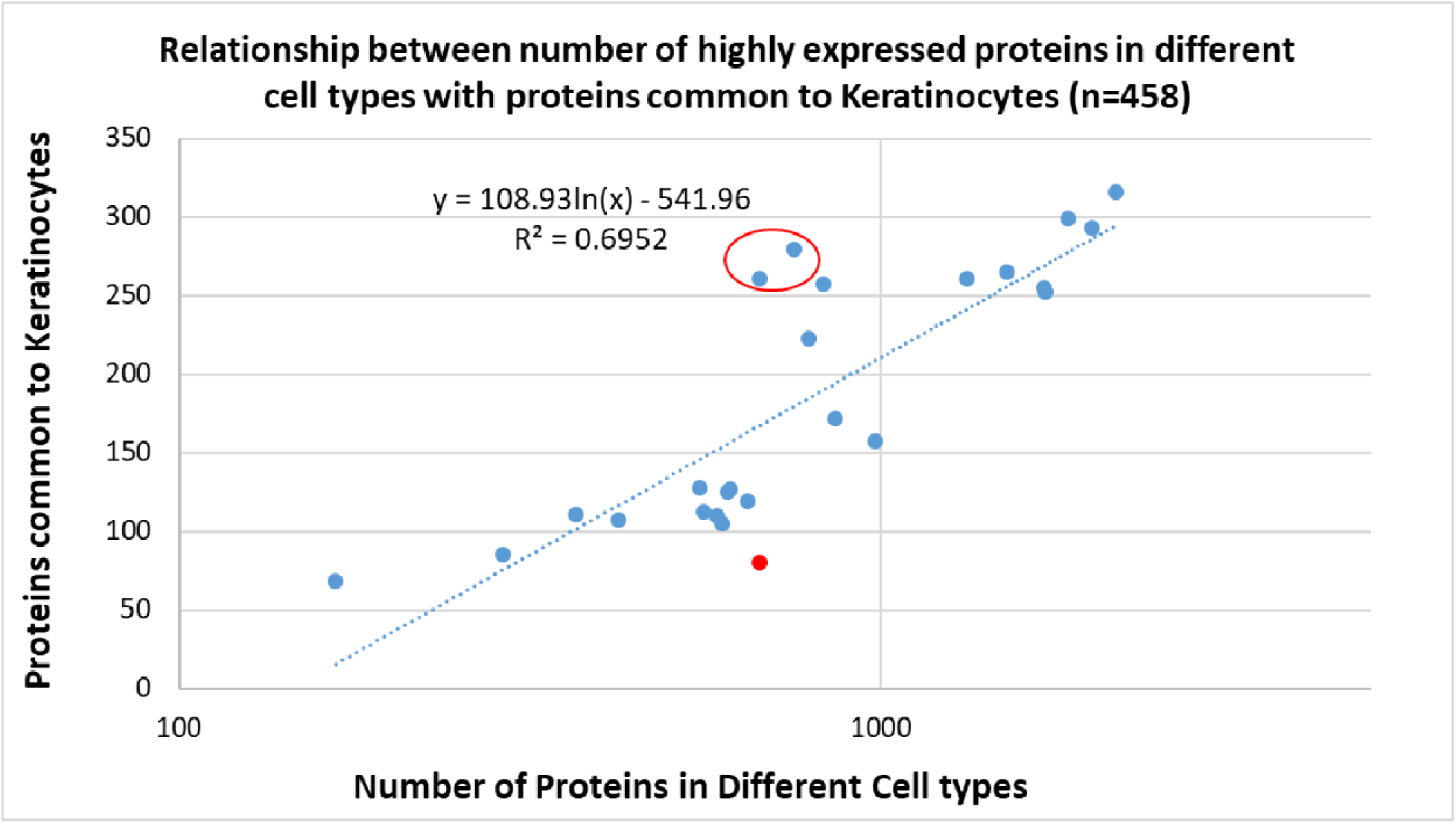
Relationship between number of highly expressed proteins in different cell types (25 cell types, with common proteins compared to keratinocytes n=>60). The trend line was depicted by y = 108.93ln(x) – 541.96 with R² value of 0.6952. Red dots represent hepatocytes and circled blue dots represent vaginal squamous epithelial cells (upper right) and cervix squamous epithelial cells (lower left).

**Figure 8:**
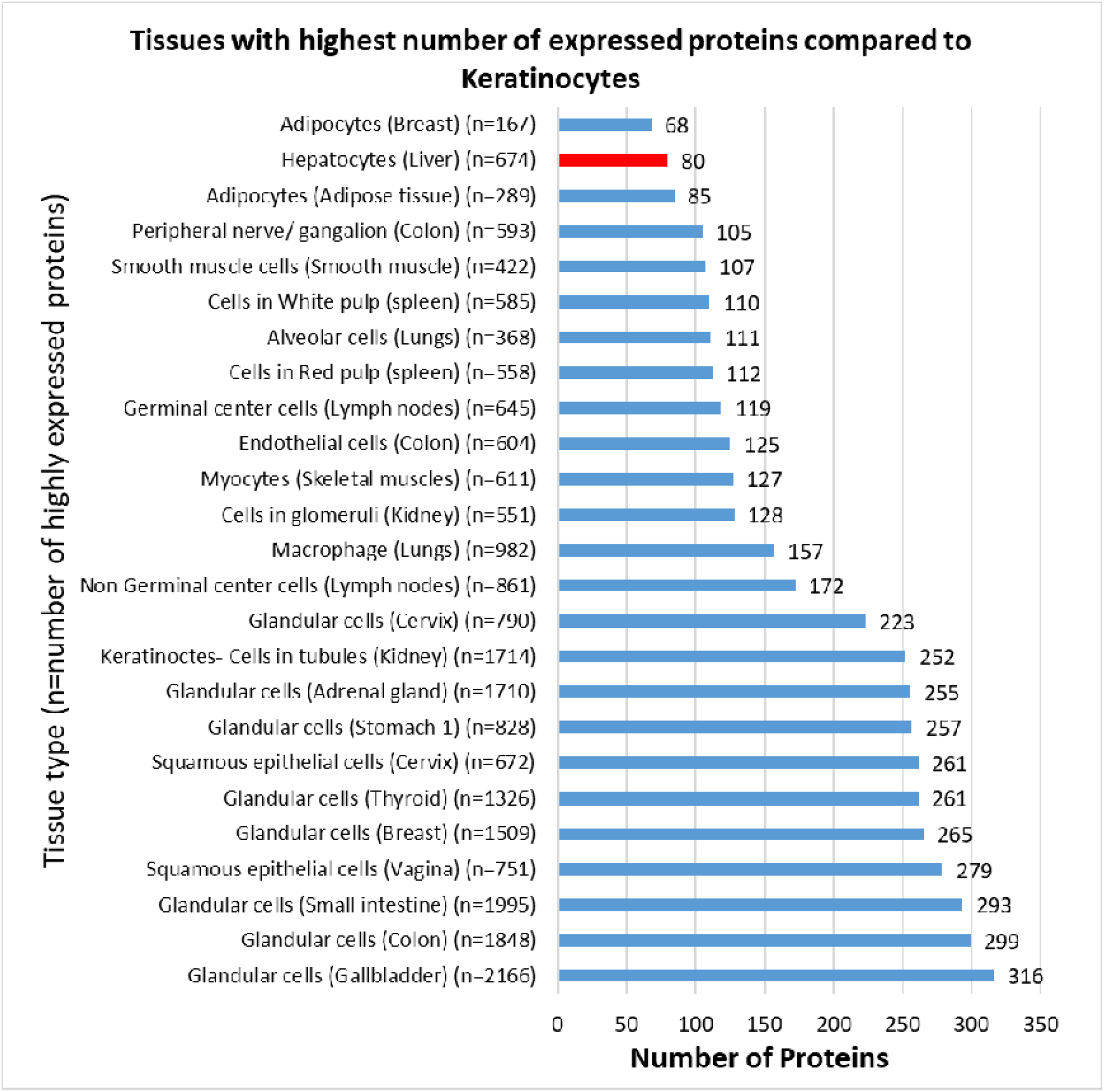
Tissues/cell types with highest number of common proteins expressed compared to keratinocytes. Only those with more than 60 (n=25 tissues/cell types) common proteins are shown.

#### Amino acid preferences in bound peptides

Previous studies have shown that positioning of the drugs like the abacavir (Illing 2012) or the carbamazepine metabolite (Simpler 2018) to the F pocket of the peptide binding groove of their respective risk allele HLA-B*57:01 and HLA-B*15:02 respectively could shift the preference of amino acid at the P9 anchor in the cellular peptides. For example, tryptophan preference was changed to leucine or isoleucine in HLA-B*57:01 and tyrosine/alanine/methionine preference was changed to glutamine and lysine in HLA-B*15:02. We checked, by in silico methods, for amino acid preferences for peptides that bound HLA-B*13:01 for proteins highly expressed in different tissues (proteins common to those expressed in keratinocytes). As mentioned above, only those tissues with at least 60 proteins (arbitrarily chosen) in common with keratinocytes were analyzed as we sought to produce significant results. We used NetMHCpan4.1 database (<u>NetMHCpan 4.1 – DTU Health Tech – Bioinformatic Services</u>) to predict the 9-amino acid long peptides compared to other online peptide prediction tools like IEDB or SYFPEITHI because only NetMHCpan4.1 had functional provision to use 4 digit resolution of HLA-B*13:01. When those with an EL score of less than 0.1 were used, the most common 9th amino acids for almost all tissues (except for cells in tubule of kidney) were leucine, phenylalanine and isoleucine. While it would have been more fruitful to see the differences in amino acid preferences at position 9 with or without dapsone, we could, due to limitation of resources, only quantitate the amino acid preferences of PBGs without dapsone. We also looked at amino acid preferences at positions 7 and 8. It could be hypothesized that even a small proportion of a suitable amino acid at position 9, 8 or 7 could be later over-represented in the presence of dapsone. But this difference should have been peculiar for hepatocytes only as SCARs are usually seen in keratinocytes and hepatocytes. We found no differences in amino acid preferences at positions 9, 8 and 7 of the tissues. The trend of amino acid preferences were very similar in all tissues including the hepatocytes. There was a slight difference in amino acid usage at position 8 for hepatocytes compared to other tissues. Glycine (G), histidine (H) and isoleucine (I) amino acid preferences were similar for hepatocytes at position 8 whereas it was usually low for G and I and slightly high for H for all other cell types/tissues. Whether this has any biological significance is not known.

**Figure 9:**
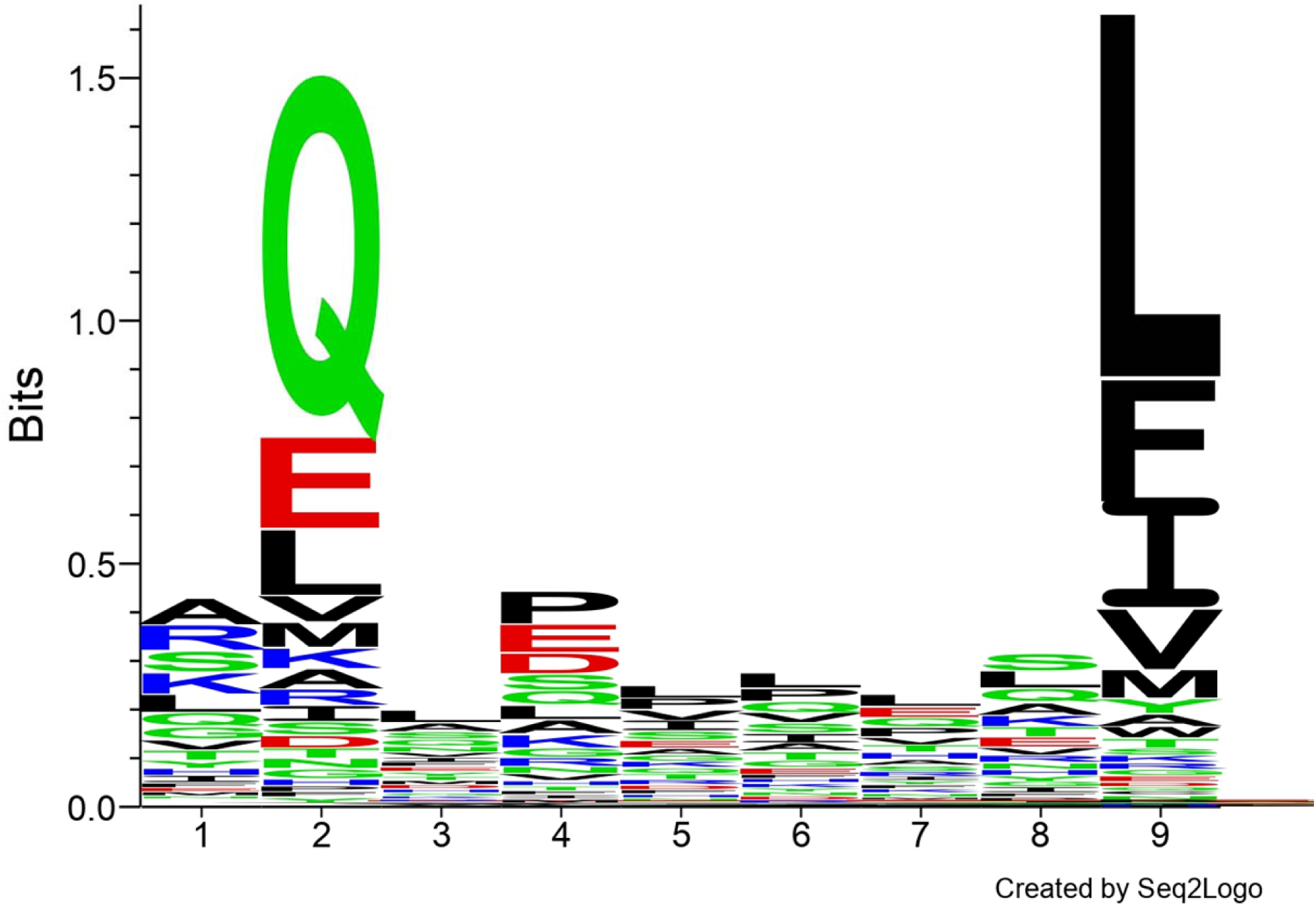
Amino acid preferences for peptides from hepatocytes calculated using Seq2logo application. Proteins for peptide prediction was selected based on commonality with the keratinocyte expressed protein (see text).

**Figure 10:**
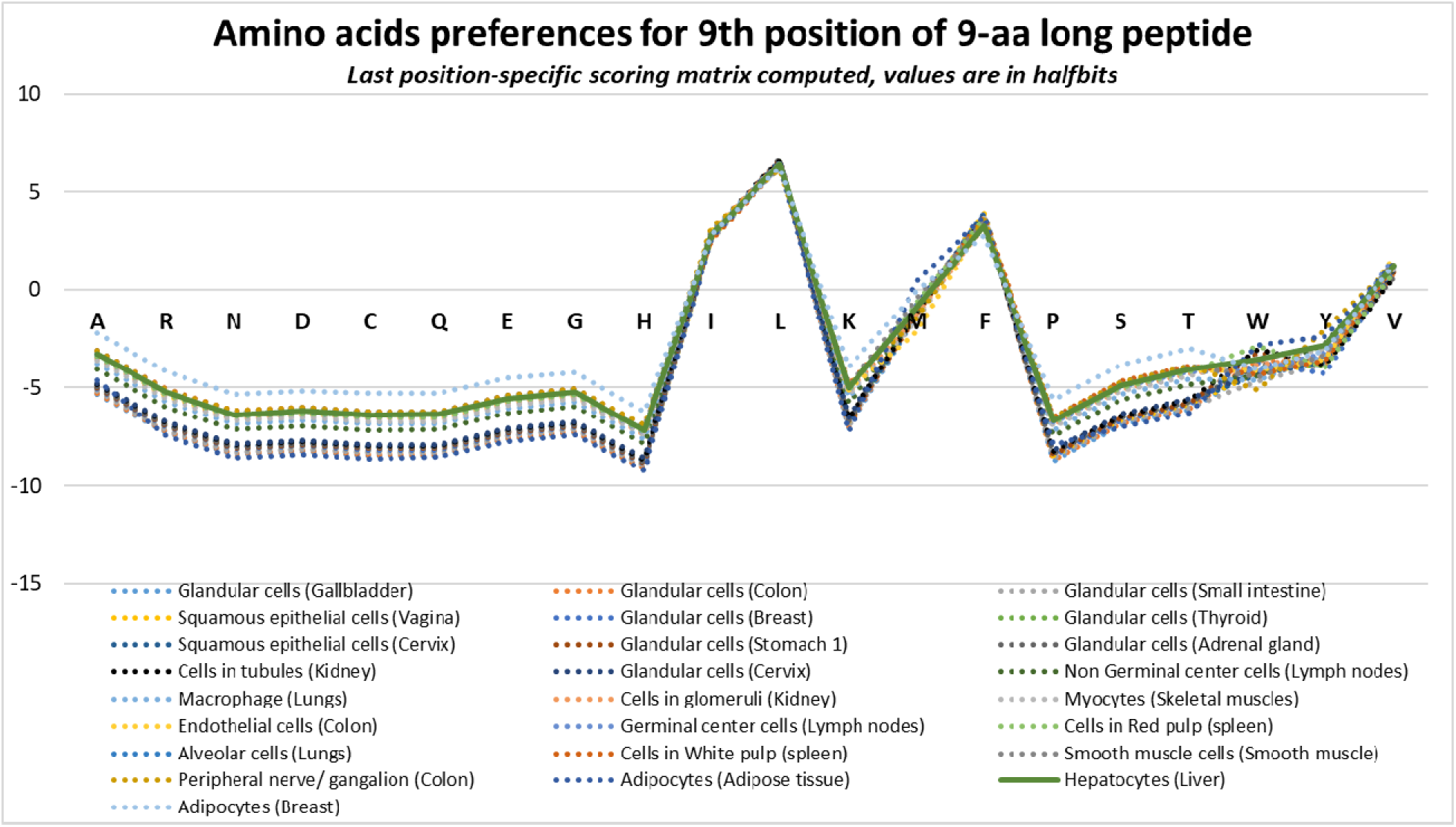
Last position-specific scoring matrix computed for amino acid preferences at 9th position calculated using Seq2logo application. All tissues are shown in dotted lines except hepatocyte in bold green.

**Figure 11:**
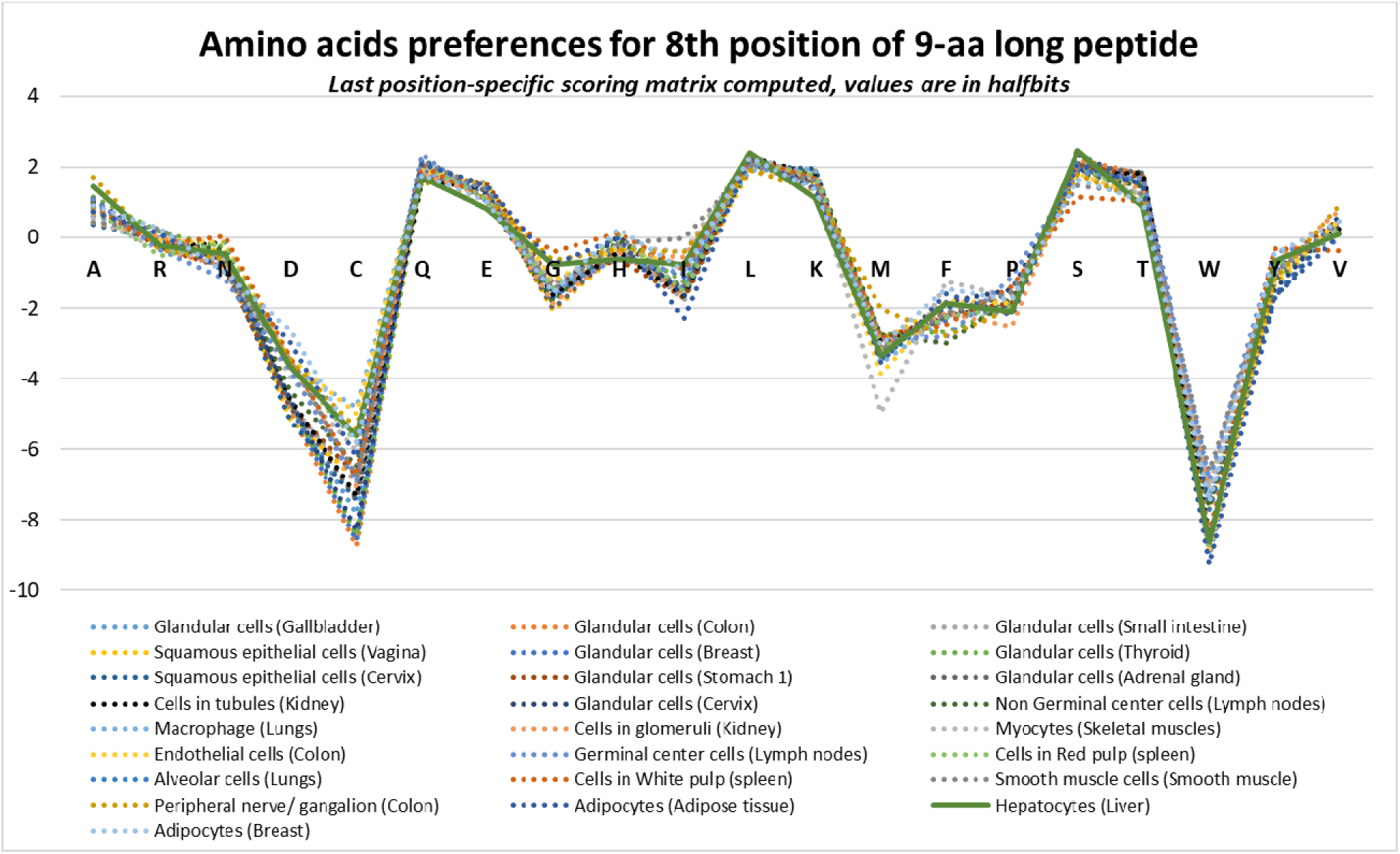
Last position-specific scoring matrix computed for amino acid preferences at 8th position calculated using Seq2logo application. All tissues are shown in dotted lines except hepatocyte in bold green.

**Figure 12:**
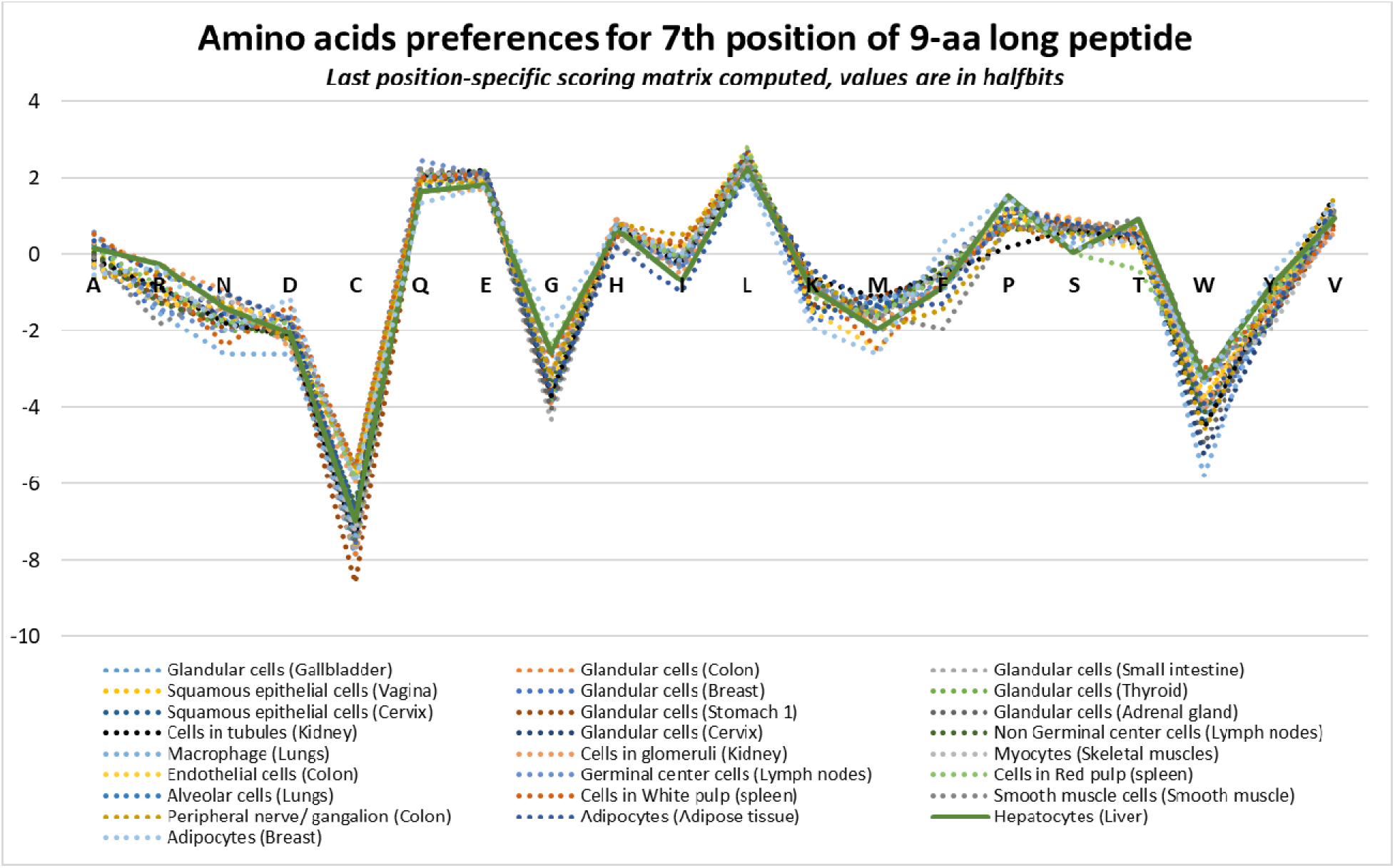
Last position-specific scoring matrix computed for amino acid preferences at 7th position calculated using Seq2logo application. All tissues are shown in dotted lines except hepatocyte in bold green.

### Gene Ontology and KEGG Pathway analysis

As no significant information was obtained based on amino acid preferences in peptides selected through HLA-B*13:01 in different tissues, GO, KEGG and Reactome analyses were performed to identify similarities in biological processes between keratinocytes and hepatocytes. Analyses were not restricted to proteins common to keratinocytes but included all proteins (“approved” and “enhanced” reliability, “highly” expressed proteins, as mentioned in the Methods section). Unfortunately, no biological processes or pathways that matched between keratinocyte and hepatocytes were identified by any of these analyses. There were 20 biological processes (GO:0002181, GO:0000075, GO:0006401, GO:0000077, GO:0031570, GO:0000288, GO:0043487, GO:0061013, GO:0043488, GO:0006402, GO:0060211, GO:0042274, GO:0016072, GO:0042254, GO:0006364, GO:1903311, GO:0044773, GO:0044774, GO:0030490 and GO:0044818) identified by GO analyses for keratinocytes but none were similar to hepatocytes. There were 178 GO pathways identified for hepatocytes. There was only one pathway for keratinocyte identified by KEGG, hsa04330, which represents Notch signalling pathway. While hepatocytes did not echo this, only two other tissues out of around 75 examined had this pathway, fibroblast in Skin 1 and squamous epithelial cells in vagina. There were 41 KEGG pathways identified in hepatocytes. There were 54 and 74 pathways identified for hepatocytes and keratinocytes by Reactome analyses. None were common in both.

#### Limitations

This study is limited by various means. Secondary data for RNA or protein expression were used for analysis. While it would have been best to study the expression of HLAs or proteins from actual drug allergic and non-allergic cases, it would have been highly resource demanding too. Thus, use of data from public databases can be useful in the sense that these data have been validated and but also, data from highly varying tissue types are available for simultaneous comparisons. For drug:HLA affinity analyses, while biochemical and immunological experiments could help provide more conclusive answer, molecular docking is still a safe preliminary task that should precede wet lab experiments. Due to resource-limitations, we used two different methods of molecular docking. Results show that both of these methods could differentiate between the known binder (HLA-B*13:01) and non-binder (HLA-B*13:02). Thus, the results obtained could be considered reliable. In spite of that, not all stable affinity of dapsone to HLA-Bs were unanimous in both models used during SWISSDOCK (2 models per allele). In this study, we used data from an alternative method (AutoDock VINA) and epidemiology, to summarize the possible HLA alleles related to dapsone binding. The alleles predicted to have high affinity with dapsone in our study are subjective and further confirmation will be required. For immunopeptidome analyses, this study could not find any differences in peptides expressed in hepatocytes compared to other tissues. This could be due to use of protein expression data at homeostatic conditions. Analyzing protein expression data during hypersensitivity might have found expressional differences.

#### Conclusion

The differential expression of HLA-B, especially its higher levels in males and older age groups, provides a plausible explanation for the epidemiological trends of SCARs. The strong expression of HLA-B in sun-exposed skin emphasizes its potential role in immune responses triggered by external stimuli. Molecular docking analyses further identified non-HLA-B13:01 alleles with significant binding affinities for dapsone, offering explanations for hypersensitivity in individuals lacking the HLA-B13:01 allele. These findings suggest that variations in HLA expression and binding affinity contribute to the variability in hypersensitivity prevalence, supporting the need for personalized approaches in predicting and managing drug-induced SCARs. We could not find similarities in GO, KEGG and Reactome pathways in liver and skin, with respect to highly expressed genes. The use of “highly expressed” genes may have limited the analysis for similarity. Or, some other factors may come into play. For example, presence of T cells with skin and liver homing receptors (Zhao 2019). Zhao et al (2019) found expression of CXCR3 and CCR4 in CD4 T cells whereas CXCR3, CCR4, CCR10, CCR9 and CCR6 in CD8 T cells. Where the T cells get primed have significant roles on where the activated T cells ultimately reach (Brinkman 2013). Both skin and liver memory T cells express α4β1 integrin which may be responsible for presence of similar T cells in these two tissues. (Fu 2016). Similarly, presence of drug metabolism pathways in both skin and liver, increasing the local tissue concentration of the culprit drugs in these tissues., could also predispose skin and liver preferentially for injuries (Line 2023). Various other approaches may identify reasons why these two organs are frequently associated with drug hypersensitivities.

## Data Availability

All data produced in the present study are available upon reasonable request to the authors

